# Symptoms of pediatric feeding disorders among individuals with 3q29 deletion syndrome

**DOI:** 10.1101/2020.09.18.20197301

**Authors:** Addam J. Wawrzonek, T. Lindsey Burrell, William Sharp, Scott E. Gillespie, Rebecca M Pollak, The Emory 3q29 Project, Melissa Murphy, Jennifer Gladys Mulle

## Abstract

**Objective:** To evaluate symptoms of pediatric feeding disorders in a sample of individuals with 3q29 Deletion Syndrome. Previous research has found that individuals with 3q29 deletion syndrome (3q29Del) may experience elevated feeding concerns in early childhood; however, the specificity of these feeding concerns in this pediatric population is not well understood.

**Methods:** We compared individuals with 3q29Del (n = 60) to matched controls (n = 59) using an 11-item survey that assessed commonly reported symptoms associated with pediatric feeding disorders. An exploratory analysis also examined individuals with 3q29Del with and without a comorbid autism spectrum disorder (ASD) diagnosis.

**Results:** Caregivers of 3q29Del cases reported higher incidences of feeding concerns on all 11 items included in the survey. This included statistically significant differences in food refusal behaviors, rejection of one or more food group, and a history of failure to thrive. Parents of children with comorbid autism were more likely to report concerns regarding rejection of one or more food group compared to children with 3q29Del without autism.

**Conclusions:** Results suggest individuals with 3q29Del experience increased symptoms of pediatric feeding disorders. Future research should include a more thorough multidisciplinary evaluation to further document the severity and identify optimal remediation strategies.

## Introduction

3q29 deletion syndrome (3q29Del) is caused by a rare 1.6 Mb heterozygous deletion on chromosome 3,^1^ with an estimated prevalence of one in 30-40,000 in the general population.^2^ The syndrome is associated with a range of neurodevelopmental and neuropsychiatric phenotypes, including mild to moderate intellectual disability (ID), increased risk for autism spectrum disorder (ASD) and/or anxiety disorders, and a 40-fold increased risk for schizophrenia.^3^ Individuals with 3q29Del also are at higher risk for failure to thrive, reduced birth weight, and significant growth deficits.^4^ In addition to evidence of ID, ASD and growth concerns, a recent systematic survey of a large 3q29Del cohort also found significantly higher rates of global developmental delays (GDD) and gastrointestinal complications compared to the general population.^1^ Together, high prevalence of these co-occurring conditions suggest individuals with 3q29Del may also be at high risk for pediatric feeding disorders.

Pediatric feeding disorders involve severe disruptions in nutritional and caloric intake exceeding ordinary variations in hunger, food preference, and/or interest in eating.(citation) Restriction in intake may involve food selectivity (i.e., eating a narrow variety of foods and/or rejecting one or more food groups) and/or food refusal (i.e., eating a restricted volume of food resulting in formula or feeding tube dependence).^9^ Individuals who severely restrict the volume and/or variety of foods consumed during meals meet diagnostic criteria for Avoidant and Restrictive Food Intake Disorder (ARFID).^10^ Manifestations of ARFID include faltering growth, nutritional deficiencies, and the need for oral nutrition supplementation or tube feeding to support growth and meet energy and nutrient needs. Many complex factors contribute to the development of this level of feeding concern, including behavioral and environmental variables, oral motor skill deficits, and/or medical or anatomical abnormalities.^12,13^ Indeed, it is well-established that there is a higher incidence of feeding disorders among individuals with ASD and GDD, as well as children with predisposing medical conditions (e.g., congenital or acquired respiratory, cardiac, and gastrointestinal [GI] problems).^5,6,7,8^ Children with ARFID and food selectivity most often present with comorbid developmental conditions including ASD and/or DD; ^7,11^ children with ARFID and formula or feeding tube dependence most often present with complex medical histories.^8,12^

Provisional evidence suggest that as many as 64% of children with 3q29Del experience elevated feeding problems during the first year of life.^1^ Available evidence, however, does not provide specific detail regarding the nature of feeding problems in this pediatric population. The purpose of the present study was two-fold. The primary aim was to evaluate symptoms of pediatric feeding disorders in a sample of individuals with 3q29Del relative to a control sample. A secondary aim was to analyze symptom presentation among individuals with 3q29Del and comorbid ASD relative to individuals with 3q29Del sample without ASD. In the process, the study provides insight into the topography and prevalence of feeding problems associated with 3q29Del.

## Methods

### Materials

Individuals with 3q29 deletion were ascertained through the existing 3q29 registry housed at Emory University (3q29.org), as previously reported.^1^ Recruitment of study subjects, informed consent, assent, HIPPA authorization, and data collection are all enabled by the website. This process is approved by Emory University’s Institutional Review Board (IRB00064133). Data collection instruments exist within the registry; answers may be submitted by an individual with 3q29 deletion syndrome or a family member (usually a parent or guardian) if the patient is underage or not capable of submitting answers. At registration, baseline information is collected about the participant (the individual with 3q29Del), including gender and birthdate, and contact information is obtained for the participant and the informant.

A custom medical and demographic questionnaire is deployed within the registry and includes seven health-related domains that were intentionally prioritized for data collection based on medical issues previously noted in 3q29 individuals.^4,17^ A prior analysis of questionnaire data revealed a high rate of feeding problems, with 64% of respondents reporting feeding problems in infancy.^1^ Additionally, anecdotal data from parents participating on the registry (JGM) and a local feeding clinic (WS) independently suggested that individuals with 3q29 deletion syndrome were susceptible to clinically significant feeding problems. In response to this we developed an eleven-item questionnaire to gather detailed data on the kinds of feeding problems experienced by study subjects with 3q29 deletion syndrome (see supplement 1: Feeding Questionnaire). This questionnaire was deployed through the registry.

### Participant Characteristics

60 3q29Del registrants (48.33% male) were included in the present study, ranging in age from 0.5-35.9 years (mean = 9.13 years). Of these participants, 15 (25%) reported an ASD diagnosis. Control study subjects were recruited via emails sent to intramural CDC and Emory Listservs that invited a community sample to fill out surveys in an identical fashion to cases. Controls reporting a clinical diagnosis of any neurodevelopmental disorder were excluded (N = 1). A total of 59 control participants (50.84% male) were included in the study. Description of the study sample can be found in Table 1.

**Table 1:** Characteristics of study participants with 3q29Del and controls

| Variable | 3q29Del<br>N=60 | Control<br>N=59 |
| --- | --- | --- |
| Age, years (mean, range) | 9.13 (0.5-35.9) | 10.5 (1.7-41.6) |
| Gender, male (*n*, percent) | 29 (48.33) | 30 (50.84) |
| Comorbid Diagnosis (*n*, percent)* |  |  |
| Autism | 15 (25) | 0 |
| Gastrointestinal Concerns | 39 (65) | 8 (13.55) |
| Global Developmental Delay | 26 (43.33) | 0 |
| Intellectual Disability | 14 (23.3) | 0 |
| None of the Above | 11 (18.3) | 51 (86.45) |

*\*Will not add up to 100% as participants could have more than one comorbid diagnosis*

### Analysis

Variables from the medical questionnaire (age at completion, gender, autism diagnosis) were coded as follows: age at completion, numerical value in years; gender, male yes/no; autism diagnosis yes/no. Variables from the feeding questionnaire (feeding tube dependency, formula dependency, little food by mouth, history of failure to thrive, picky eater, smooth food only, does not feed self, history of anemia, food refusal behaviors, and total food group refusal) were coded as follows: feeding tube dependency, yes/no, formula dependency, yes/no, little food by mouth, yes/no, history of failure to thrive, yes/no, picky eater, yes/no, smooth food only, yes/no, does not feed self, yes/no, history of anemia, yes/no. Total food group refusal was coded as a count, ranging 0-4 for total refusal of the following four food groups; fruits, vegetables, proteins and starches. Food refusal behaviors was coded as a count, ranging 0-8 for the following eight reported food refusal behaviors during meals; crying, pushing away the spoon, disruptions (defined as removing food or utensils from the table), spitting out food, aggression, leaving the table, negative statements, and other).

We first summarized continuous and discrete variables in a univariate fashion, comparing cases to controls using t-tests for continuous data and chi-squre or Fisher’s exact test for discreet data. We then performed multivariate analysis, adjusting for age and gender. Specifically, total food group refusals and food refusal behaviors were assessed using negative binomial regression, assessing each outcome as a count. Rate ratios were calculated both unadjusted and adjusted for confounders (i.e., age and gender). Remaining outcomes variables were dichotomous (yes versus no; e.g., Picky Eater, Tube Dependency) and analyzed using binary logistic regression. Odds ratios are shown unadjusted and adjusted for confounders, with 95% confidence intervals and adjusted ratio p-values. When expected frequency counts were less than 5, exact binary logistic regression was employed; moreover, when odds ratios could not be calculated due to 0 observations in a 2×2 contingency table cell, Fisher’s exact p-values were presented. To assess the effect of ASD as a potential comorbidity in 3q29 participants, a sub-analysis considered only 3q29Del study subjects, comparing self-report ASD diagnosis to those without a self-reported ASD diagnosis. Statistical models for this sub-analysis were as described above. All analyses were performed in SAS v.9.4 (Cary, NC).

## Results

### Feeding concerns in 3q29Del versus controls

Reported feeding concerns within the 3q29Del cohort (N=60) were compared to the control cohort (N=59), while controlling for age and gender (Table 2). Individuals with 3q29Del reported significantly higher rates of food refusal behaviors and refusal of all foods from one or more food groups. Specifically, the rate of food refusal behaviors in 3q29Del participants was 17.1 times higher (Mean ± SD: 1.9 ± 2.1) than that of controls (Mean ± SD: 0.1 ± 0.5). Similarly, the rate of number of food groups refused in 3q29Del participants was 3.81 times higher (Mean ± SD: 0.9 ± 1.2) than that of controls (Mean ± SD: 0.2 ± 0.6). Participant responses also indicated significantly higher odds of 3q29Del participants being reported as a picky eater (OR=8.30; CI=3.50, 19.7), inability to feed themselves (OR=14.5; CI=1.81, 116.6), and a history of anemia (OR=19.0; CI=3.85, 93.9), relative to Control participants.

**Table 2:**
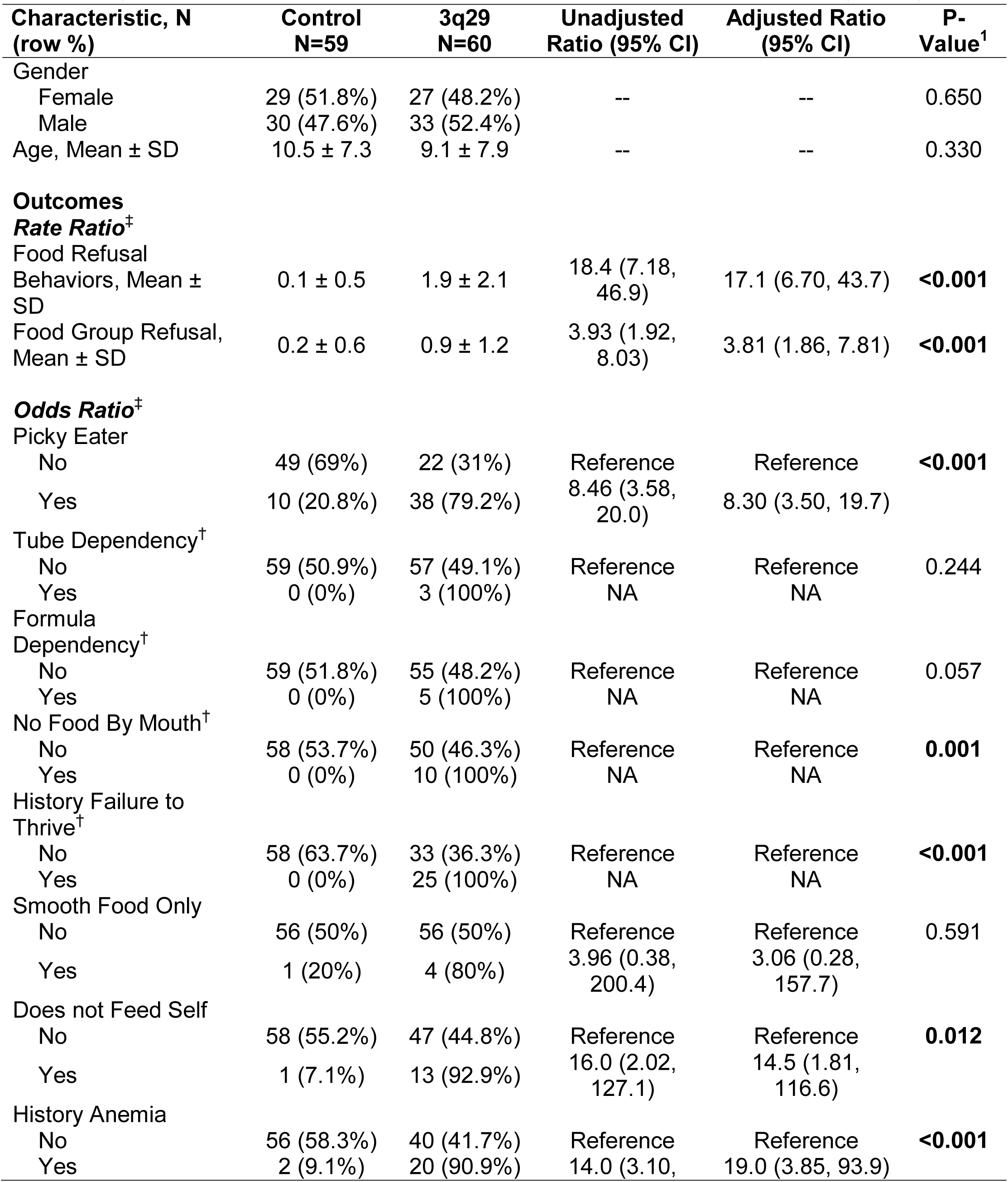
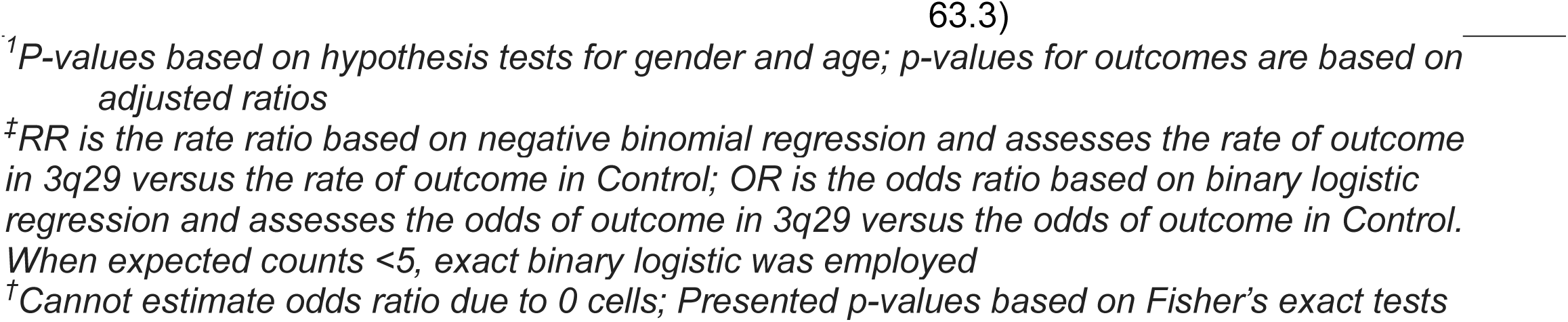
Adjusted risks and odds of outcomes for 3q29 patients vs. control patients (N=119)

Additionally, there were a number of outcome variables (i.e., tube dependency, formula dependency, no food by mouth and history of failure to thrive) that indicated a proportion above 0% in 3q29Del participants, but the controls had no cases. When this occurred, only summary statistics and exact p-values are provided. Of these variables for the 3q29Del group, 3 (5% vs. 0%, p=0.244) reported feeding tube dependency, 5 (8.3% vs. 0%, p=0.057) reported formula dependency, 10 (16.7% vs. 0%, p=0.001) reported accepting little to no food by mouth, and 25 (41.7% vs. 0%, p<0.001) reported a history of failure to thrive (Table 2).

### Feeding concerns in 3q29Del with and without comorbid ASD

We next performed a sub-analysis of individuals with 3q29Del, comparing a comorbid autism diagnosis (N=15) to those without an autism diagnosis (N=45), while controlling for age and gender (Table 3). There was a significant difference for age, where individuals with a comorbid ASD diagnosis were 4.8 years older on average (p=0.041). The data indicated a significantly higher number of food groups refused for individuals with 3q29Del and comorbid ASD compared to those without ASD. The rate of number of food groups refused in participants with 3q29Del and comorbid ASD was 2.28 times higher (Mean ± SD: 1.3 ± 1.2) than those without comorbid ASD (Mean ± SD: 0.7 ± 1.1).

**Table 3:** Adjusted risks and odds of outcomes for 3q29 patients, ASD+ versus ASD- (N=60)

| Characteristic, N<br>(row %) | Without<br>ASD<br>N=45 | With ASD<br>N=15 | Unadjusted<br>Ratio (95% CI) | Adjusted Ratio<br>(95% CI) | P-Value<sup>1</sup> |
| --- | --- | --- | --- | --- | --- |
| Gender |  |  |  |  |  |
| Female | 22 (81.5%) | 5 (18.5%) | -- | -- | 0.294 |
| Male | 23 (69.7%) | 10 (30.3%) |  |  |  |
| Age, Mean ± SD | 7.9 ± 7.4 | 12.7 ± 8.3 | -- | -- | **0.041** |
| **Outcomes** |  |  |  |  |  |
| **Rate Ratio<sup>†</sup>** |  |  |  |  |  |
| Food Refusal Behaviors, Mean ± SD | 1.8 ± 1.9 | 2.1 ± 2.7 | 1.20 (0.55, 2.63) | 1.65 (0.73, 3.76) | 0.225 |
| Food Group Refusal, Mean ± SD | 0.7 ± 1.1 | 1.3 ± 1.2 | 1.88 (0.87, 4.04) | 2.28 (1.02, 5.11) | **0.049** |
| **Odds Ratio<sup>†</sup>** |  |  |  |  |  |
| Picky Eater |  |  |  |  |  |
| No | 17 (77.3%) | 5 (22.7%) | Reference | Reference |  |
| Yes | 28 (73.7%) | 10 (26.3%) | 1.21 (0.36, 4.16) | 1.54 (0.41, 5.85) | 0.524 |
| Tube Dependency<sup>†</sup> |  |  |  |  |  |
| No | 42 (73.7%) | 15 (26.3%) | Reference | Reference | 0.566 |
| Yes | 3 (100%) | 0 (0%) | NA | NA |  |
| Formula Dependency<sup>†</sup> |  |  |  |  |  |
| No | 40 (72.7%) | 15 (27.3%) | Reference | Reference | 0.318 |
| Yes | 5 (100%) | 0 (0%) | NA | NA |  |
| No Food By Mouth<sup>†</sup> |  |  |  |  |  |
| No | 36 (72%) | 14 (28%) | Reference | Reference | 1.000 |
| Yes | 9 (90%) | 1 (10%) | 0.29 (0.01, 2.46) | 0.53 (0.01, 6.90) |  |
| History Failure to Thrive<sup>†</sup> |  |  |  |  |  |
| No | 26 (78.8%) | 7 (21.2%) | Reference | Reference | 0.230 |
| Yes | 17 (68%) | 8 (32%) | 1.75 (0.54, 5.71) | 2.20 (0.61, 7.99) |  |
| Smooth Food Only |  |  |  |  |  |
| No | 43 (76.8%) | 13 (23.2%) | Reference | Reference | 0.167 |
| Yes | 2 (50%) | 2 (50%) | 3.23 (0.22, 48.7) | 3.36 (0.43, NA)* |  |
| Does not Feed Self |  |  |  |  |  |
| No | 36 (76.6%) | 11 (23.4%) | Reference | Reference | 0.289 |
| Yes | 9 (69.2%) | 4 (30.8%) | 1.45 (0.27, 6.57) | 3.36 (0.47, 27.9) |  |
| History Anemia |  |  |  |  |  |
| No | 30 (75%) | 10 (25%) | Reference | Reference | 0.957 |
| Yes | 15 (75%) | 5 (25%) | 1.00 (0.29, 3.45) | 1.04 (0.25, 4.37) |  |

<sup>1</sup>P-values based on hypothesis tests for gender and age; p-values for outcomes are based on adjusted ratios
<sup>‡</sup>RR is the rate ratio based on negative binomial regression and assesses the rate of outcome in 3q29 versus the rate of outcome in Control; OR is the odds ratio based on binary logistic regression and assesses the odds of outcome in 3q29 versus the odds of outcome in Control. When expected counts <5, exact binary logistic was employed
<sup>†</sup>Cannot estimate odds ratio due to 0 cells; Presented p-values based on Fisher's exact tests
<sup>\*</sup>Upper bound could not be estimated due to few stratified observations within the adjusting covariates

Although the only statistically significant finding when examining individuals with 3q29Del and comorbid ASD relative to those without ASD was food group refusal, a number of other variables indicated clear clinically meaningful findings, but were likely not statistically significant due to reduced statistical power as a result of a low sample size of individuals with comorbid ASD. The rate of food refusal behaviors for individuals with comorbid ASD was 1.65 times higher than those without ASD. Likewise, the odds of individuals with comorbid ASD would be described as picky eaters were 1.54 times higher, the odds that they would have a history of failure to thrive were 2.2 times higher, the odds that they would only eat smooth/puree foods were 3.36 times higher, and the odds that they would not feed themselves were 3.36 times higher than those without comorbid ASD. While exploratory, these moderate and large effects indicate that individuals with comorbid ASD are at a much greater risk for pediatric feeding disorders and the affiliated medical complications, relative to those without ASD, in our cohort.

## Discussion

Previous research has found that individuals with 3q29Del report significant challenges with feeding in the first years of life.^1^ The purpose of the present study was to conduct a preliminary analysis of the specific behavioral and nutritional concerns related to pediatric feeding disorders within this population relative to a control sample. The initial data indicates that a significantly higher proportion of individuals with 3q29Del reported symptoms related to pediatric feeding disorders. These individuals reported higher rates of total refusal of one or more food groups, and increased food refusal behaviors during mealtimes. Of note, reported risk for each variable were substantial; individuals with 3q29Del were 17 times more likely to report food refusal behaviors, including crying, negative statements, throwing food and utensils, spitting food, elopement and aggression (figure 1). Similarly, rates of total refusal of all foods from one or more food groups were 8.3 times higher in this population, with 40% of participants refusing all food from at least one food group (figure 2). Finally, individuals with 3q29Del reported higher rates of tube and formula dependence, poorer self-feeding skills, dependence on puree foods to meet their needs, and were more likely to have a history of anemia or failure to thrive.

**Figure 1:**
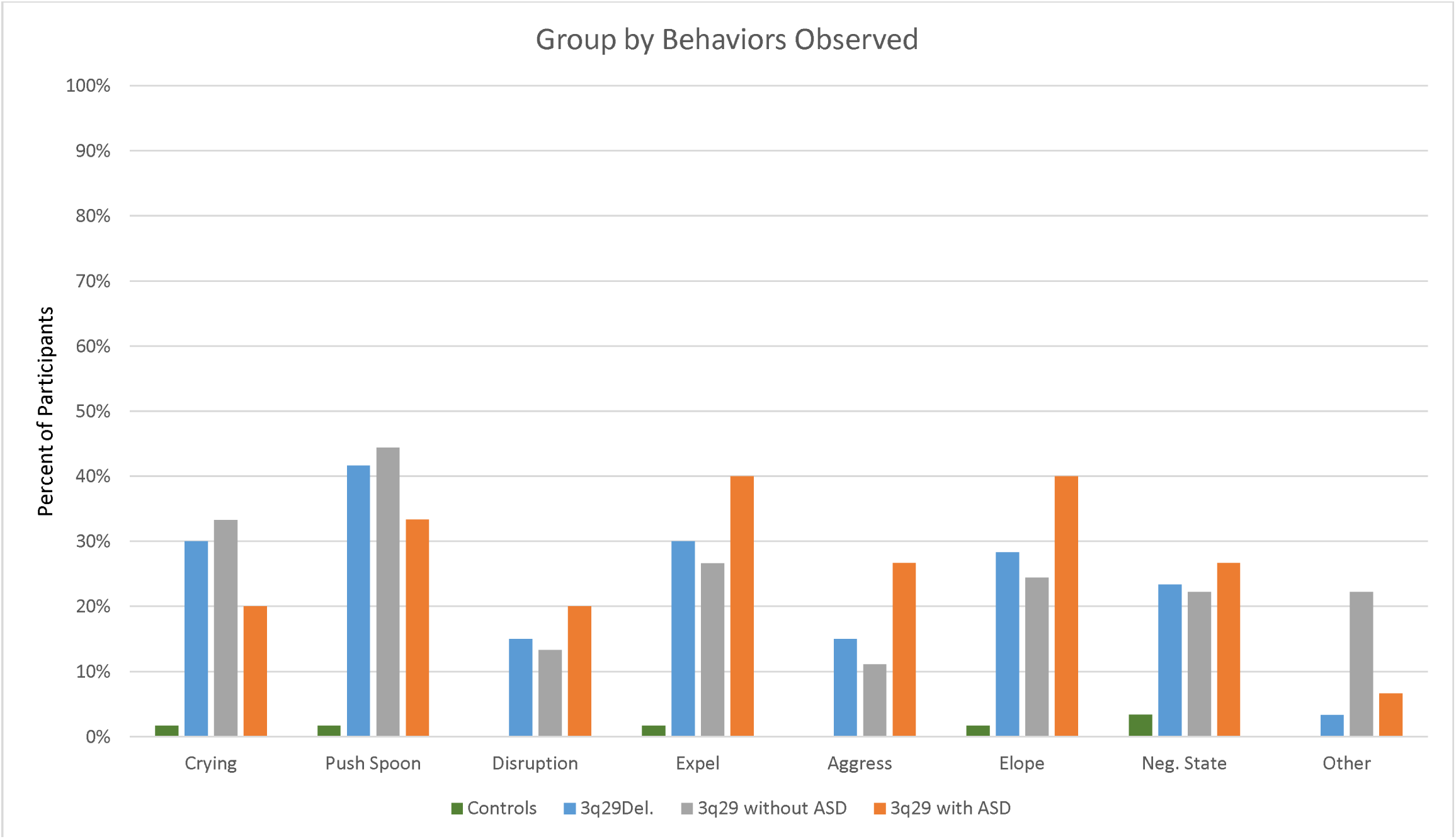
Food refusal behavior by group

**Figure 2:**
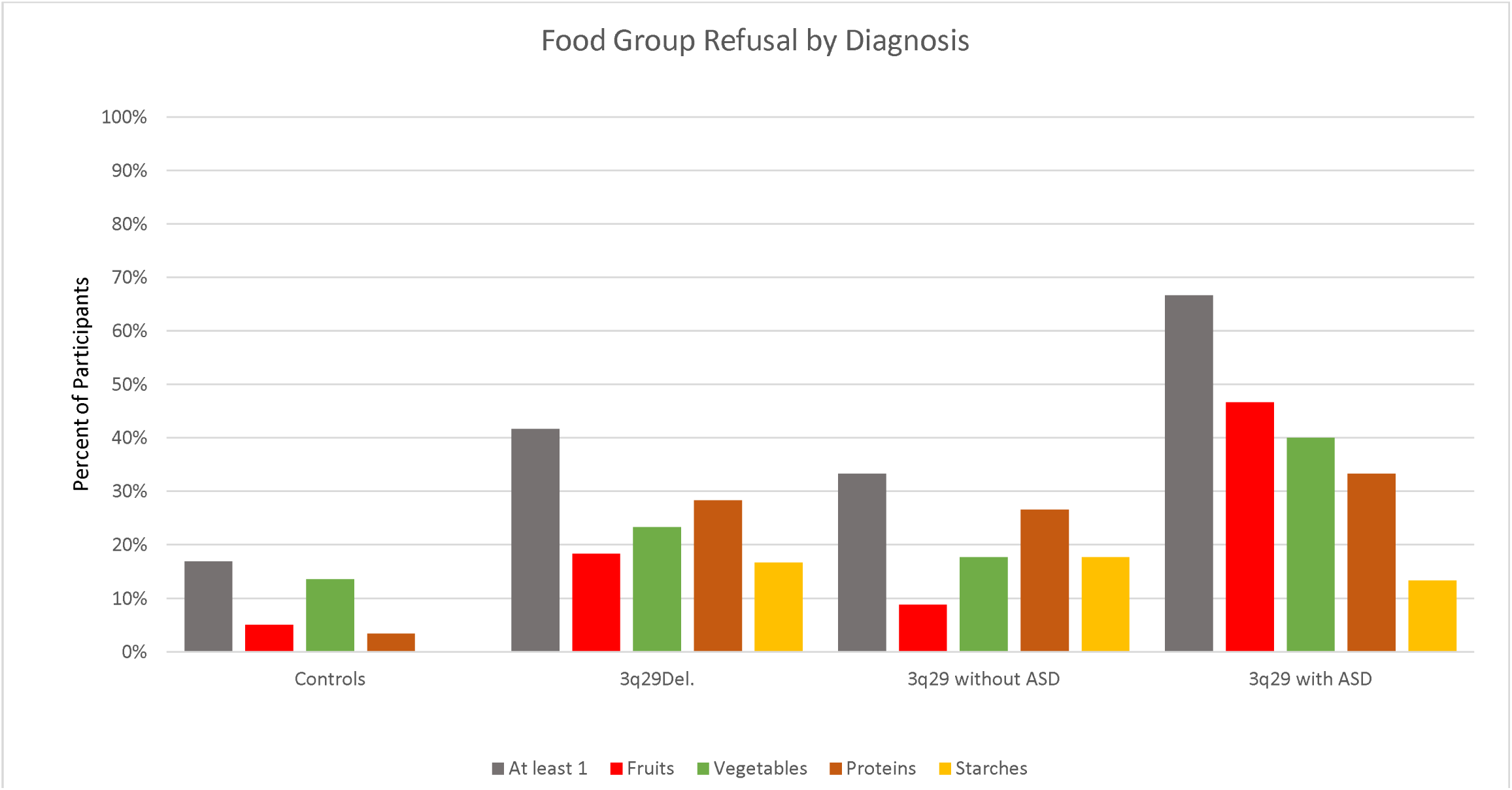
Total refusal of all foods from a food group by diagnosis

Secondly, pediatric feeding disorders occur much more frequently in children with autism spectrum disorder,^5,7^ which is a common comorbid diagnosis within the 3q29Del population.^1^ Understanding how feeding concerns differ across individuals in this population with and without autism spectrum disorder can help practitioners better identify those with the most significant risk within the entire 3q29Del population. The data indicated that, within the 3q29Del sample, the rate of refusal for all foods from one or more food groups was 2.29 times higher for individuals with 3q29Del who also had a comorbid diagnosis of autism. This group also showed clinically elevated levels (increase of 50% or more) of refusal behaviors during meals (table 2), as well as increased odds of being described as a picky eater, dependency on tube feedings, formula or puree foods, poor self-feeding skills, and a history or failure to thrive and/or anemia. Within specific reported feeding behaviors, individuals with 3q29Del reported higher rates of all behaviors relative to controls, but those with comorbid ASD reported higher rates across five out of eight categories relative to those without comorbid ASD (figure 1). Similarly, 33.3% of individuals with 3q29Del but without autism refused all food from at least one food group (compared to 16.9% of controls), whereas 66.67% of individuals with comorbid ASD reported refusing all foods from at least one food group (figure 2). These data together indicate that all individuals with 3q29Del are likely to report behavioral and medical concerns related to pediatric feeding disorders, but those with comorbid ASD are at a much greater risk.

### Implications

The present study has a number of important implications for children with 3q29Del. Severe food selectivity as reported here places children at significant risk for nutritional deficiencies that can impact physiological and cognitive development.^16^ Individuals with 3q29Del are already at increased risk for neurodevelopmental deficits. Micronutrient deficiency, insufficient caloric intake and poor growth can have significant impacts on cognitive development and so can exacerbate the neurodevelopmental concerns in this population. Children presenting with feeding problems also frequently experience compromised immune functioning, may require recurrent hospitalizations, or in the most severe cases, the placement of medically costly and invasive nasogastric or gastrostomy tubes to maintain caloric and nutritional intake.^18,19^ Additionally, parents of children with pediatric feeding disorders report high rates of emotional distress,^20^ which can directly affect the socioemotional functioning of caregivers and children,^21,22^ and cause disruption in the overall parent-child relationship.^23,24^

Given the cognitive, developmental, and psychosocial implications of pediatric feeding disorders for individuals with 3q29Del, it is essential that screening for pediatric feeding concerns be a part of the standard assessment protocol for children within this population. A number of evidence based practices have been well established for increasing oral intake of foods for children that can reestablish feeding practices which serve to meet their caloric and nutritional needs on a daily basis. Current best practice follows an intensive, multidisciplinary intervention (IMI) in day hospital settings to expand oral food intake and reduce dependence formula or enteral nutrition supplementation.^14^ A systematic review of these practices found an average 70% success rate for a full wean from enteral feeding and a 74% increase in oral consumption of foods.^8^ Early intervention and increase of oral food consumption can help prevent or reduce the negative impacts of poor nutrition on childhood growth and development, and therefore should be a high priority for pediatricians working with children with 3q29Del.

### Limitations

The present study had a number of important limitations to consider. First, all data used in was collected via self-report from parent’s and caregivers of children with 3q29Del. Self-report data has the potential to have poor reliability and validity.^25^ Additionally, any diagnostic, developmental or medical data was also collected via self-report, as opposed to coming from a standardized assessment by an appropriate trained professional. With regards to pediatric feeding concerns, no data was collected on the intensity or frequency on reported behaviors, and so it cannot be determined how disruptive the reported behaviors were to mealtimes. Additionally, although the concerns related to pediatric feeding were more specific than in previous studies, no direct, clinical assessments were conducted to determine if any concerns were clinically significant enough to warrant a diagnosis. Furthermore, no direct oral motor assessments were conducted to assess for oral motor skill deficits, nor were there direct nutritional assessments conducted to evaluate disruption to dietary intake. Statistical power was also an issue, as the autism spectrum disorder subgroup only had 13 participants. While this subgroup reported clinically significant elevations in most feeding concerns, most did not meet the threshold for statistical significance. Finally, the individuals in the control group were screened out for neurodevelopmental disorders, which are associated with pediatric feeding concerns. A more true comparison between 3q29Del and the general population would include a sample of these disorders as well.

### Future Directions

This study was the first to specifically evaluate pediatric feeding concerns within the 3q29Del population, and provides directions for future research in a number of key areas. Subsequent studies should conduct more thorough, direct assessments of feeding issues to determine the severity and prevalence of feeding concerns which cannot be evaluated by self-report alone. Additionally, more direct data on nutritional deficiencies as well as oral motor skill development would help to clarify the underlying concerns related to pediatric feeding disorders. Finally, 3q29Del is comorbid with several other neurodevelopmental and medical disorders which are associated with pediatric feeding concerns, including intellectual disability, developmental delay, and various gastrointestinal conditions.^5,6,7,8^ More information is needed on the prevalence of pediatric feeding concerns within specific subgroups and how they relate to the greater 3q29Del population.

## Conclusion

The present study found that Individuals with 3q29Del report a number of concerns related to pediatric feeding disorders, including significant disruptive mealtime behaviors, total refusal of all foods from one or more food groups, picky eating, poor self-feeding skills, and a history of anemia and failure to thrive. Individuals with a comorbid diagnosis of autism present with similar or higher rates of the above concerns. Given that this population is already at risk for developmental and physiological deficits and delays, it is essential that feeding problems be identified and addressed in early childhood. Early treatment of malnutrition or poor caloric intake can soften the impact on development. Now that these concerns have been identified within this population, future research should evaluate specific nutritional, oral motor and behavioral problems to identify the severity of pediatric feeding disorders.

## Data Availability

De-identified data will be made available to qualified investigators with appropriate CITI training.

